# First positronium image of the human brain *in vivo*

**DOI:** 10.1101/2024.02.01.23299028

**Authors:** P. Moskal, J. Baran, S. Bass, J. Choiński, N. Chug, C. Curceanu, E. Czerwiński, M. Dadgar, M. Das, K. Dulski, K.V. Eliyan, K. Fronczewska, A. Gajos, K. Kacprzak, M. Kajetanowicz, T. Kaplanoglu, Ł. Kapłon, K. Klimaszewski, M. Kobylecka, G. Korcyl, T. Kozik, W. Krzemień, K. Kubat, D. Kumar, J. Kunikowska, J. Mączewska, W. Migdał, G. Moskal, W. Mryka, S. Niedźwiecki, S. Parzych, E. Perez del Rio, L. Raczyński, S. Sharma, Shivani, R.Y. Shopa, M. Silarski, M. Skurzok, F. Tayefi, K. Tayefi, P. Tanty, W. Wiślicki, L. Królicki, E. Ł. Stępień

## Abstract

Positronium, an unstable atom consisting of an electron and a positron, is abundantly produced within the molecular voids of a patient’s body during positron emission tomography (PET) diagnosis. Its properties, such as its average lifetime between formation and annihilation into photons, dynamically respond to the submolecular architecture of the tissue and the partial pressure of oxygen molecules. However, the diagnostic information that positronium may deliver about early molecular alterations remains unavailable in clinics with state-of-the-art PET scanners.

This study presents the first *in vivo* images of positronium lifetime in humans. We developed a dedicated J-PET system with multiphoton detection capability for imaging. The measurements of positronium lifetime were performed on a patient with a glioblastoma tumor in the brain. The patient was injected intratumorally with the ^68^Ga radionuclide attached to Substance-P, which accumulates in glioma cells, and intravenously with ^68^Ga attached to the PSMA-11 ligand, which is selective to glioma cells and salivary glands. The ^68^Ga radionuclide is routinely used in PET for detecting radiopharmaceutical accumulation and was applied for positronium imaging because it can emit an additional prompt gamma. The prompt gamma enables the determination of the time of positronium formation, while the photons from positronium annihilation were used to reconstruct the place and time of its decay. The determined positronium mean lifetime in glioblastoma cells is shorter than in salivary glands, which in turn is shorter than in healthy brain tissues, demonstrating for the first time that positronium imaging can be used to diagnose disease *in vivo*. This study also demonstrates that if current total-body PET systems were equipped with multiphoton detection capability and the ^44^Sc radionuclide was applied, it would be possible to perform positronium imaging at 6500 times greater sensitivity than achieved in this research. Therefore, it is anticipated that positronium imaging has the potential to bring a new quality of cancer diagnosis in clinics.

## I. INTRODUCTION

Positronium is abundantly produced in the patient’s body during positron emission tomography (PET) and its properties (e.g. mean lifetime) are modified by the tissue sub-molecular environment [1, 2]. We have invented a method to image positronium lifetime [3–5] based on the registration of two photons from its annihilation and additional prompt gamma emitted by some of the isotopes (e.g. ^68^Ga routinely used in PET diagnostics [6–8]). The method was recently validated in *ex vivo* studies by reconstructing positronium images [9] and by observing differences in positronium mean lifetime between healthy and tumor tissues [10–13]. Here we demonstrate the first *in vivo* positronium images of a human brain performed with a dedicated PET scanner designed to enable a simultaneous registration of annihilation photons and prompt gamma.

In a standard PET scan, biomolecules (tracers) labeled with a radionuclide that emits positrons are administered to the patient at picomolar concentration. In the patient’s body, positrons annihilate with electrons creating photons whose registration in the PET scanner enables imaging of the distribution of the labeled biomolecules in the body. Therefore, PET allows for the assessment of metabolic effects, transport activities, or receptor expression *in vivo* directly in the patient.

PET diagnostics has experienced continuous improvements in radiation detection [14] and radiopharmacy [7], from the first blurry brain tumor images achieved with two crystal scintillation detectors in 1953 [15], through the first application of fluorodeoxyglucose labeled with ^18^F for brain metabolism imaging in 1979 [16] and the first scans with ^68^Ga labeled somatostatin receptors in 1999 [6], to the dynamic and simultaneous imaging of all patient tissues demonstrated in 2019 [17] with the total-body PET system constructed from 564,480 crystal scintillators [18].

However, after 70 years of development, the main principle of PET scanners of recording electron-positron annihilation into two photons and its applications based on localizing the accumulation of pharmaceuticals remained unchanged. For example, PET scans of the brain are mainly performed to locate lesions, including determining the location and size of brain tumors [19], to assess the measurement of cellular and tissue metabolism, to show blood flow, to evaluate patients with seizure disorders (epilepsy) who do not respond to pharmacotherapy [20], and patients with memory impairment due to neurodegenerative diseases (Alzheimer’s disease spectrum and Parkinson’s disease [21]). The detection of molecular alterations which is not currently achievable would be possible using positronium imaging, which would increase PET specificity for early recognition of these diseases before the functional changes occur [1].

In the body, a positron emitted from the isotope attached to the biomolecule may annihilate with an electron either directly (*∼*60% cases) or via the formation of positronium (*∼*40% cases) [2]. Positronium is a metastable atom made of an electron and a positron. It may be formed as a long-lived 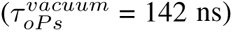 spin-one ortho-positronium (oPs), or as a short-lived 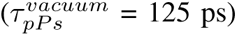 spin zero para-positronium (pPs). In vacuum, oPs decays into 3-photons (oPs *→* 3γ) and pPs into 2-photons (pPs *→* 2γ) [2]. In the tissue (Fig. 1) the oPs lifetime 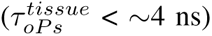 is significantly shortened, since in addition to self-annihilation into 3-photons, the positron from oPs may pick off electrons from the surrounding molecular environment (pick-off process) and annihilate predominantly into 2-photons. The oPs lifetime may also be shortened by interacting with the dissolved oxygen molecules. oPs may react with a paramagnetic O_2_ molecule *via* spin-exchange or oxidation processes [22, 23]. In the spin-exchange process, oPs is converted to pPs, which then rapidly decays into two photons (oPs + O_2_ *→* pPs + O_2_ *→* 2γ + O_2_). Therefore, the smaller the inter- and intra-molecular voids and the higher the oxygen concentration, the shorter the mean ortho-positronium lifetime. Fig. 1 indicates possible decay mechanisms of positronium in the intra-molecular spaces.

**Fig. 1:**
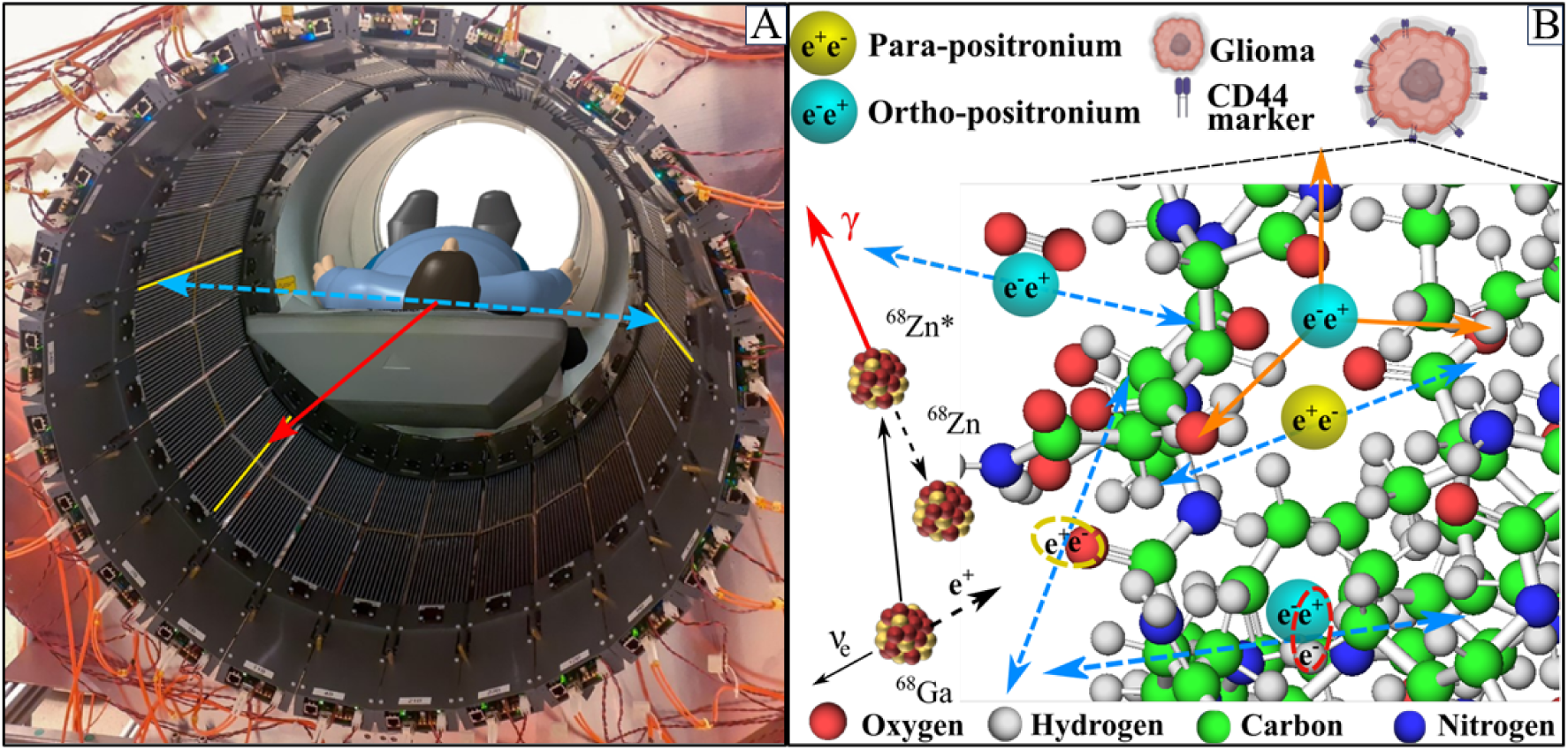
**(A)** Photograph of the patient in the modular J-PET tomograph taken during the first positronium imaging of a human brain. The patient diagnosed with primary brain glioma was intravenously and intratumorally administered with pharmaceuticals labeled with the ^68^Ga radionuclide, which emits positrons and prompt gamma rays (the course of imaging is illustrated in Fig. 2). The superimposed arrows represent photons from electron-positron annihilation (blue-dashed) and prompt gamma from the deexcitation of the ^68^Zn* radionuclide (red-solid). The tomograph consists of 24 modules each built from 13 plastic scintillator strips. These plastic strips in which the gamma rays interacted have been highlighted in yellow. **(B)** Pictorial illustration of the ^68^Ga isotope decay chain (^68^Ga *→* ^68^Zn* + e^+^ + ν *→* ^68^Zn + γ + e^+^ + ν) and possible ways of positron and positronium annihilation within molecules on the example of the PSMA molecule in the receptor overexpressed at the membrane of the glioma cell. Positron (black-dashed) emitted by ^68^Ga may annihilate directly (e^+^e*^−^ →* photons) or via formation of positronium (e^+^e*^−^ →* Ps *→* photons) [1]. Positronium (Ps) may be formed as para-positronium (pPs, yellow) or ortho-positronium (oPs, blue). Blue-dashed arrows indicate annihilations into two photons used for imaging. Main annihilation mechanisms mentioned in order of decreasing probabilities include [2]: (i) direct annihilation (yellow ellipse), (ii) annihilation of a positron from ortho-positronium by picking off the electron from the atom (red ellipse), (iii) self-annihilation of para-positronium inside free space between atoms, (iv) conversion of oPs to pPs on O_2_ molecule proceeded by self-annihilation of pPs (oPs + O_2_ *→* pPs + O_2_ *→* 2γ + O_2_), and (v) self-decay of oPs into three photons (orange arrows) inside free space between atoms. The distances and size of atoms are shown to scale with the diameter of positronium twice as large as hydrogen [2].

Current PET scanners utilize e^+^e*^−^* annihilation into 2-photons only, and they are insensitive to the positron annihilation mechanism which may deliver diagnostics information complementary to the presently available images of metabolic rate and receptor expression. For example, the early tissue alterations at the molecular level and the changes in the tissue oxidation could be sensed by knowing the ratio of 3γ to 2γ decay rate and the mean positronium lifetime [1, 2, 23]. Therefore positronium is considered as a promising biomarker of tissue pathology at the early stage of molecular alterations and as a biomarker of tissue oxidation [2, 23, 24].

Here, we demonstrate for the first time the *in vivo* images of positronium lifetime in the human brain. Fig. 1 shows the photograph of the patient in the modular J-PET tomograph. The superimposed arrows indicate annihilation photons (dashed blue) from the electron-positron annihilations, while red solid arrow shows the prompt gamma from the ^68^Zn isotope deexcitation. In order to determine the positronium lifetime in the patient, it is necessary to know the time of positronium formation and the time of its annihilation. The moment of annihilation is determined from the time when annihilation photons (blue dashed arrows in Fig. 1) interact in the detection system. The time of the positronium formation may be estimated if the radionuclide, in addition to the positron, emits also a prompt gamma (red arrow in Fig. 1). Registering the time when prompt gamma interacts in the detector system enables us to estimate the time of positronium formation [4, 5].

In this study, the patient diagnosed with primary brain glioma was administered pharmaceuticals labeled with ^68^Ga radionuclide that in 1.3% of positron emissions emits also a prompt gamma (Fig. 1). Imaging of the patient was performed using a dedicated novel modular J-PET scanner, equipped with the acquisition system enabling simultaneous registration of signals from annihilation photons and prompt gamma. In the presented research we applied the newly introduced positronium imaging method [3, 4] that was recently successfully validated in the first *ex vivo* positronium imaging of phantoms comprising cardiac myxoma tumor and healthy adipose tissues operated from patients [9]. The observed dependence of the mean positronium lifetime on the tissues type [10, 12, 25–27], and on the grade of cancer malignancy [28] encourages to perform *in vivo* positronium imaging in order to test the hypothesis if positronium may serve as a parameter in the *in vivo* diagnosis in clinics. This study presents the first-ever clinical positronium lifetime images and a discussion of the prospects of its application in clinical imaging with total-body PET scanners.

### II. RESULTS

The main result of the presented research is the demonstration of the first *in vivo* image of positronium lifetime in the human head. The research was performed in such a way as not to interfere with routine diagnostics and therapy. The examined patient was a man in his 40s, with a recurrent secondary glioblastoma (II/IV) in the right frontoparietal lobe. The primary aim of the conducted therapy was to destroy the glioma tumor using alpha particles emitted by the ^225^Ac radionuclide. The demonstration of positronium imaging was possible thanks to the concurrent theranostic application of the ^68^Ga isotope to monitor the site of cancer lesions using a PET scanner. The study protocol, approved by the Ethical Committee of the Medical University of Warsaw, is presented pictorially in Fig. 2A indicating the diagnosis and treatment workflow. The location and the stage of the glioblastoma tumor were determined by administering a radio-pharmaceutical [^68^Ga]-PSMA-11 to the patient in a dose of 178 MBq [8, 19]. PSMA is a Prostate Specific Membrane Antigen having high affinity to the receptors at the cell membrane of glioblastoma cancer [8]. After 66 minutes, the standard PET/CT images were acquired using a PET/CT Biograph 64 TruePoint scanner. Subsequently, the patient was moved on the table to the J-PET tomograph for a 10-minute data acquisition. The patient’s examination is documented in the photographs in Fig. 2(C-F). Subsequently, using a catheter connected to a subcutaneous port implemented into a postoperative cavity [29] (see MRI image in Fig. 3B), the patient was injected intracavitary with 20 MBq of [^225^Ac]-labeled Substance-P ([^225^Ac]Ac-DOTAGA-SP) together with [^68^Ga]-labeled Substance-P ([^68^Ga]Ga-DOTA-SP) at the dose of 8 MBq. Substance-P having high affinity to the NK-1 receptors (overexpressed on the glioblastoma cells surface and within the tumor neovasculature) was used to deliver alpha particle emitter ^255^Ac to the tumor cells [30]. The therapy was performed as previously optimized and described in reference [29], and the ^225^Ac dose activity was chosen as a compromise between the therapeutic effect and the side reaction for the treatment [31]. The [^68^Ga]Ga-DOTA-SP pharmaceutical labeled with ^68^Ga radionuclide was used to monitor the accumulation sites of the Substance-P with PET/CT Biograph 64 TruePoint. The obtained PET/CT images show the highest accumulation of [^68^Ga]Ga-DOTA-SP pharmaceutical in the cavity of the tumor (Figs. 3A, 3B, and 3C) confirming the recurrence of the glioblastoma. The images indicate also accumulation in the salivary glands since they possess receptors at the cell membranes having high affinity to the PSMA [8]. As the last step, the patient was moved on the table to the modular J-PET tomograph for the 13-minute data acquisition for positronium imaging.

**Fig. 2:**
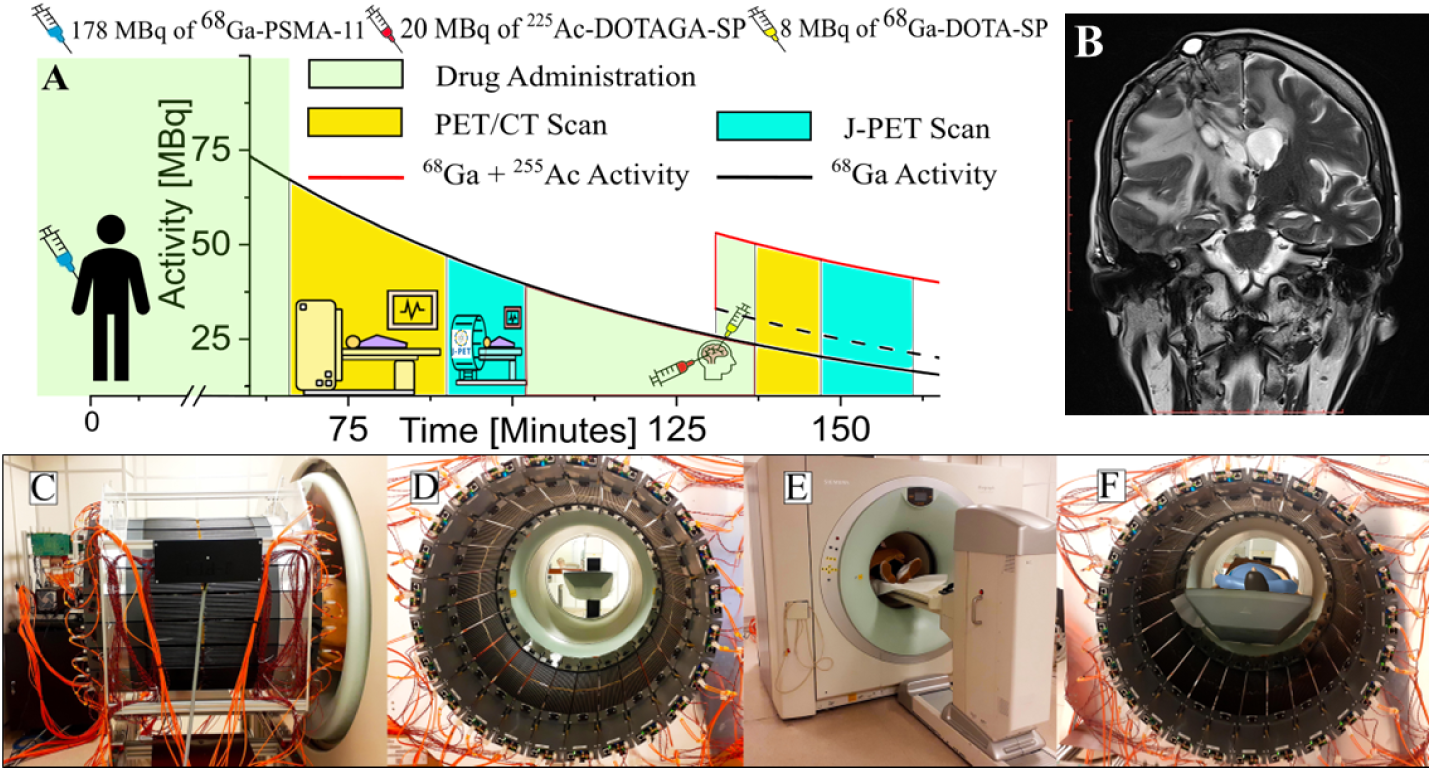
**(A)** The course of diagnosis and treatment of a patient with secondary recurrent glioblastoma. The solid black curve indicates the decrease in the activity of ^68^Ga radionuclide after intravenous injection of a 178 MBq activity of [^68^Ga]Ga-PSMA-11 radiopharmaceutical followed 131 minutes later by intratumoral administration of 8 MBq of [^68^Ga]Ga-DOTA-SP (dashed-black curve) together with the 20 MBq of [^225^Ac]Ac-DOTAGA-SP (red curve). After the first and the second administration of pharmaceuticals, the patient was imaged with the Siemens PET/CT Biograph 64 TruePoint, and then with a modular J-PET for the time indicated in the graph in yellow and turquoise, respectively. **(B)** The T2-weighed MRI coronal image of the head (Magnetom 3T, Siemens Healthcare) showing the tumor and the position of the cat-cath system (visible on the upper left part of the head) for the administration of a radiopharmaceutical for the local treatment of glioblastoma. **(C-F)** Photographs illustrating the course of patient imaging with the modular J-PET scanner. **(C)** Modular J-PET scanner placed behind the PET/CT Biograph 64. **(D)** View of the patient’s bed from the inside of the J-PET tomograph. **(E)** The patient was moved on the table so that his torso and legs were in the Biograph PET/CT and his head in the J-PET scanner. In this view, only the patient’s feet are visible on the edge of the Biograph PET/CT. **(F)** Photograph of the patient with the head inside of the J-PET scanner during imaging after intravenous administration of [^68^Ga]Ga-PSMA-11 radiopharmaceutical.

**Fig. 3:**
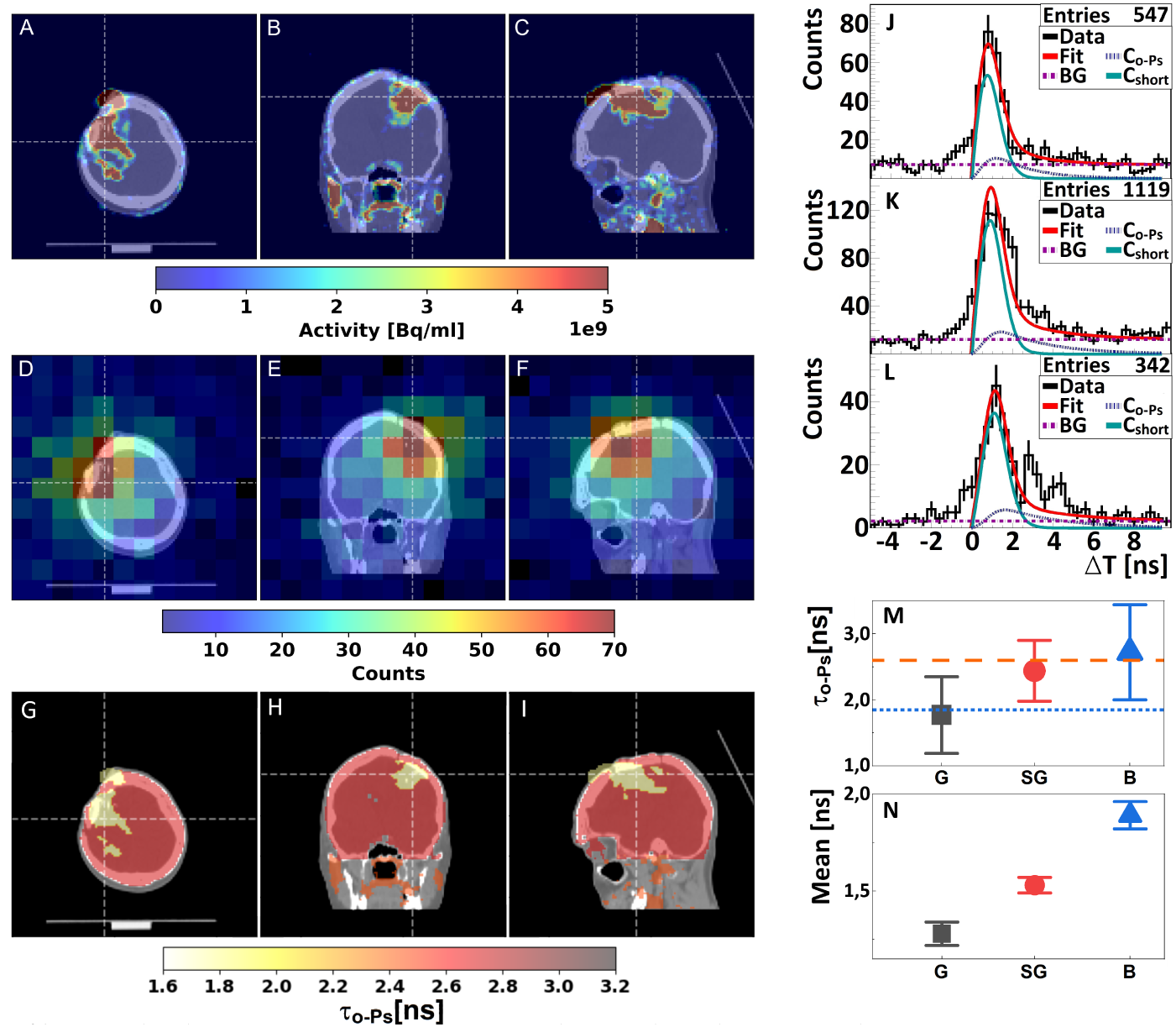
**(A,B,C)** Standard PET/CT fusion images obtained with Biograph 64 TruePoint (Siemens Medical Solutions) of the head of a patient with recurrent secondary glioblastoma in the right frontoparietal lobe. Images were acquired after intracavitary injection of the [^68^Ga]Ga-DOTA-SP together with [^225^Ac]Ac-DOTA-SP radiopharmaceuticals (imaging time course is shown in Fig. 2). An accumulation of [^68^Ga]Ga-DOTA-SP in the capsule and cavity of the tumor in transverse (A), coronal (B), and sagittal (C) planes is presented. **(D,E,F)** Images of the density distribution of positron annihilation accompanied by the emission of the prompt gamma are shown in transverse (D), coronal (E), and sagittal (F) planes. These images were acquired with the modular J-PET system soon after the regular diagnosis with Biograph 64 TruePoint (see Fig. 2). The presented spatial distribution of positronium accumulation in the brain is superimposed on the CT images. The white thin dashed lines in each (A-F) image indicate a cross-section with the other two planes presented. **(G,H,I)** Positronium images are shown in transverse (G), coronal (H), and sagittal (I) planes. **(J,K,L)** Distributions of positron annihilation lifetime (ΔT) determined in glioblastoma tumor (J), salivary glands (K), and healthy brain tissues (L). Black histograms denote experimental data. The superimposed curves indicate the result of the fit of the function describing contributions from the annihilation of ortho-positronium (C*_oPs_*), para-positronium and direct annihilations (C*_short_*), and background due to accidental coincidences (BG). The red curve denotes the sum of all contributions (Fit). The details of the fit are described in the Methods section. **(M,N)** Mean oPs lifetime (τ*_oPs_*, fig. M) and mean value of the positron lifetime in the range between 0 and 5 ns (Mean, fig. N) determined for the glioblastoma tumor (G, black squares), salivary glands (SG, red circles), and healthy brain tissues (B, blue triangles). For comparison, the blue dotted line and orange dashed line indicate the mean oPs lifetime in Cardiac Myxoma tumor (1.92 ns) and adipose tissue (2.72 ns) as determined in the *ex vivo* studies [10].

A dedicated modular J-PET scanner is built from 24 detection modules each constructed from plastic scintillators and programmable electronics [32–34]. The scanner, seen in photographs in Fig. 1A and Fig. 2C-E is described in detail in the Methods section. The data were collected using a data acquisition system, that we developed, which enabled triggerless signal acquisition [35]. The continuous triggerless data recording allowed us to collect multi-photon coincidences. Recorded data were analyzed using a dedicated J-PET programming framework [36]. The data analysis procedures, background subtraction, and positronium image reconstruction are described in detail in the Methods section. Therefore, it will only be mentioned briefly in the following.

In the first step of the analysis, the data were decoded from electronics to timestamp format, and then the recorded timestamps were used to reconstruct the registered signals. In the next step of the low-level data processing, signals were used to reconstruct the time and position of each gamma photon interaction in the tomograph. Further, the signals from annihilation photons and prompt gamma were disentangled and grouped into events. An event is referred to as the entire sequence of processes occurring in the patient’s body and the tomograph as a result of the decay of a single radionuclide. For each event, comprising two annihilation photons and prompt gamma, the annihilation place and time as well as the positron emission time were reconstructed. The set of such selected events (after the suppression of the instrumental and physical background) was used to reconstruct the images of the density distribution of annihilation places associated with the emission of the prompt gamma (Figs. 3D, 3E, and 3F).

Due to the low fraction of such events (only 1.3% in the case of ^68^Ga (Fig. 4B)) the images are characterized by low statistics. Therefore, in order to determine the positronium lifetime, the reconstructed image of the head of the patient was divided only into three regions of interest: the range around the glioma cancer (G), the healthy brain tissues (B), and the area around the salivary glands (SG). The lifetime spectra determined for these regions are shown in Fig. 3J, 3K, and 3L, respectively.

**Fig. 4:**
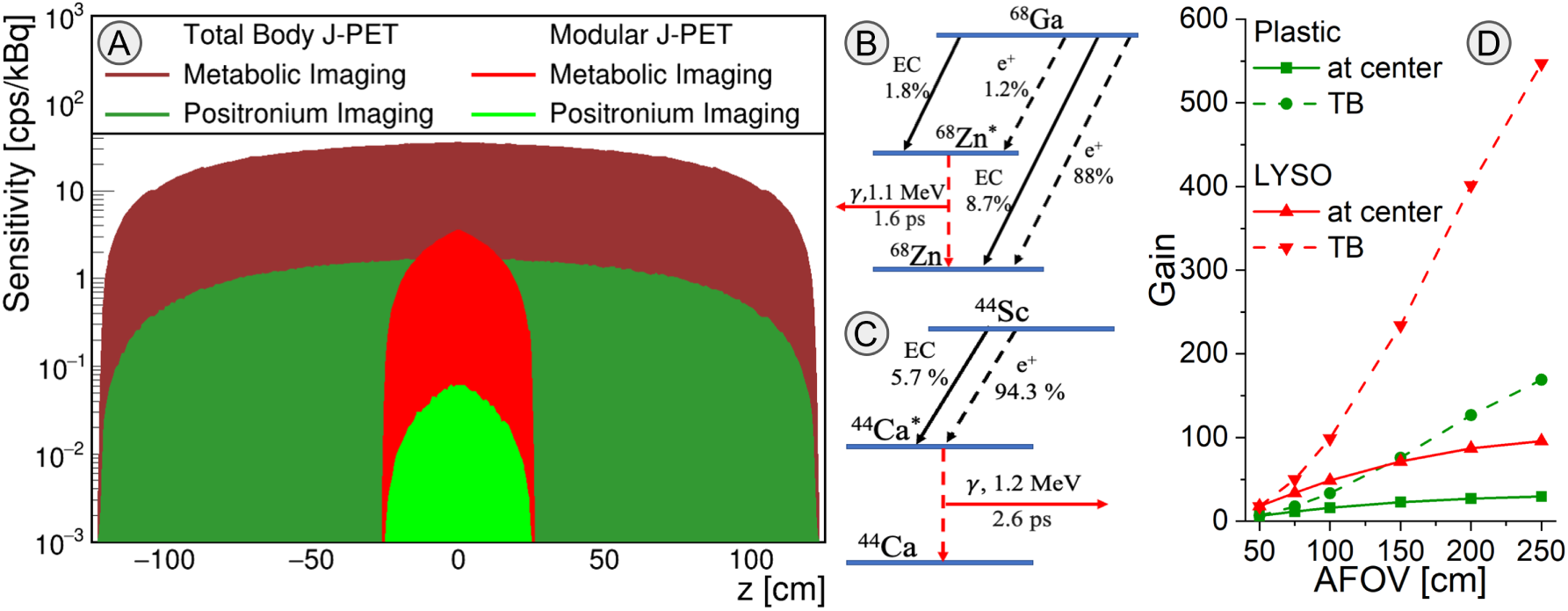
**(A)** Comparison of positronium imaging sensitivity profiles (for registration of γγ + γ*_p_* events) with sensitivity profiles for standard PET metabolic imaging (for registration of γγ events). The sensitivity for positronium imaging (green profiles) is more than a factor of ten smaller compared to the sensitivity of standard PET imaging (red profiles). This is due to the fact that positronium imaging requires the registration of prompt gamma (γ*_p_*) in addition to annihilation photons (γγ). Sensitivity profiles of (applied in this study) modular J-PET with an axial field-of-view of 50 cm, are compared to the sensitivity profiles for the design of the total-body J-PET with almost 250 cm axial field-of-view [40, 41]. **(B)**: Decay scheme of ^68^Ga radionuclide. EC denotes the electron capture process while “e^+^” denotes the emission of a positron. The mean lifetime of the excited nucleus ^68^Zn*^∗^* and the energy of the deexcitation (prompt) gamma is indicated. In this research, the ^68^Ga *→* ^68^Zn*^∗^* + e^+^ + ν *→* ^68^Zn + γ*_p_* + e^+^ + ν decay chain was used for positronium imaging which occurs only in about 1.3% (1.2/(88+1.2) *∼* 1.3) of all e^+^ emissions. **(C)** Decay scheme of ^44^Sc radionuclide, the most suited isotope for positronium imaging [41–43]. The emission of positron by ^44^Sc radionuclide is accompanied by the prompt gamma in 100% cases via ^44^Sc *→* ^44^Ca*^∗^* + e^+^ + ν *→* ^44^Ca + γ*_p_* + e^+^ + ν decay chain. **(D)** Estimated gain of positronium imaging sensitivity by increasing the axial field-of-view with respect to the modular J-PET used in this research. The gain is calculated for the sensitivity at the center (solid lines) and for the whole-body positronium imaging (dashed lines). Results for standard crystal-based PET systems (based on the uExplorer) are shown in red, and in green the result for the J-PET based on plastic scintillators is shown. The result indicates that the sensitivity at the center for positronium imaging would increase by a factor of 28 for the total-body J-PET, and by a factor of 87 for uExplorer with AFOV = 194 cm [18].

The superimposed violet-dotted curve indicates contribution due to the annihilation intermediated by ortho-positronium. Possible annihilation mechanisms are depicted in Fig. 1B. The contributions from fast annihilation mechanisms (pPs *→* 2γ and direct e+e-*→* 2γ) and annihilations via ortho-positronium (oPs *→* 2γ) were established by deconvoluting the experimental lifetime spectra by using the dedicated PALS-Avalanche software [37–39]. Details of the fitting procedure are described in the Methods section.

The determined mean ortho-positronium lifetimes (τ*_oPs_*) are presented in Fig. 3M. One observes a trend in the data. The measured value of the mean oPs lifetime in the glioma tumor (1.77 *±* 0.58 ns) falls below the salivary glands (2.44 *±* 0.46 ns), which in turn is below the measurement in healthy brain tissues (2.72 *±* 0.72 ns). Notably, the differences in the mean positron lifetimes shown in Fig. 3N are more statistically pronounced. The mean positron lifetime values were calculated (using histograms in Figs. 3J,3K,3L) in the signal range between 0 ns and 5 ns. The determined values are equal to 1.28 *±* 0.06 ns for glioma tumor, 1.53 *±* 0.04 ns for salivary glands, and 1.89 *±* 0.06 ns for healthy brain tissues. This positron lifetime measurement in healthy brain tissue is 7 standard deviations greater than the value found in glioma tumor and 5 standard deviations greater than the value for salivary glands. Combining information about the annihilations density distributions (Figs. 3A, 3B, 3C), reconstructed density distributions of e^+^e*^−^* annihilations associated with the prompt gamma emission (Figs. 3D, 3E, 3F), and the mean ortho-positronium lifetimes (Fig. 3M), the positronium lifetime image of the examined patient is determined as shown in Figs. 3G, 3H, 3I. Details are described in the Methods section.

## III. DISCUSSION AND CONCLUSIONS

This manuscript presents the first *in vivo* demonstration of positronium imaging. Fig. 3 displays images of the ortho-positronium mean lifetime in the brain of a patient with a glioma tumor. The images were acquired using the ^68^Ga radionuclide attached to the PSMA-11 and Substance-P molecules, which exhibit high affinity for receptors on the glioma cell surface. The registration of photons from positronium annihilation was employed to reconstruct the location and time of positronium decay, while the prompt gamma emitted by the ^68^Ga isotope was used to determine the time of positronium formation. This *in vivo* imaging was made possible by the specially designed and constructed multi-photon J-PET scanner, which facilitates the simultaneous registration and identification of annihilation photons and prompt gamma rays. The J-PET scanner utilized in this study was constructed as a lightweight (60 kg), modular, and portable system, enabling its straightforward installation (taking less than 1 minute) around the patient’s bed.

The mean ortho-positronium lifetime observed in healthy brain tissues (2.72 ns *±* 0.72 ns) differs from that observed in glioma tumors (1.77 ns *±* 0.58 ns), suggesting the potential of positronium imaging to enhance the specificity of PET diagnosis in *in vivo* tissue pathology assessment. While presently a one standard deviation effect, the *in vivo* determined mean ortho-positronium lifetime in glioma tumors is comparable to the previously determined *ex vivo* lifetime in cardiac myxoma tumors (1.92 *±* 0.02 ns), while the mean ortho-positronium lifetime in the healthy brain is similar to the *ex vivo* values obtained for healthy adipose tissues (2.72 *±* 0.05 ns) [9, 10].

The uncertainties in this first *in vivo* investigation are substantial due to the limited number of registered events (e.g., only 86 ortho-positronium annihilations were identified in the glioma). However, it is crucial to emphasize that the sensitivity of positronium imaging can be substantially enhanced compared to the modular J-PET employed in this study by utilizing total-body PET scanners with an extended axial field of view (AFOV). Fig. 4D shows that the sensitivity for positronium imaging will increase by a factor of 28 for the total-body J-PET, and by a factor of 87 for uExplorer with AFOV = 194 cm [18]. Based on this figure we can also roughly estimate that it will increase by a factor of about 50 for Siemens Quadra with AFOV = 106 cm [44], and by a factor of about 70 for uPennPET Explorer with AFOV = 142 cm [45]. Two of the mentioned total-body PET systems (Siemens Quadra [46] and uPenn Explorer [45]) possess data acquisition capabilities that would enable positronium imaging. Moreover, the effective sensitivity can also be significantly increased by applying other β^+^γ emitters than ^68^Ga, such as ^14^O, ^44^Sc, ^82^Rb, or ^124^I [41, 43, 47]. Of these, the use of ^44^Sc would best improve imaging efficiency as it emits the prompt gamma 100% of the time after positron emission compared to only 1.34% for ^68^Ga [41]. The decay scheme of ^68^Ga and ^44^Sc are shown in Fig. 4B and 4C, respectively. ^44^Sc radionuclide has already been applied in phantom [48] and preclinical PET imaging [49] using folates [50, 51], somatostatin analogs [52], and PSMA ligands [53, 54]. It can be produced using accelerators [54–56], or using the ^44^Ti/^44^Sc generator [57]. The application of the ^44^Sc radionuclide and total-body PET systems would enable positronium imaging with thousands of times greater sensitivity compared to the first images presented in this work (Figure 3). The sensitivity will increase by approximately 3700, 5200, or 6500 times when using Siemens Quadra, uPenn Explorer, or uExplorer, respectively. This will enable the acquisition of millions of events with coincident registration of annihilation photons and prompt gamma rays, a number comparable to standard PET imaging. Such high event statistics will enable the effective application of recently developed iterative positron imaging reconstruction methods [58–61], which can improve the spatial resolution of positronium lifetime images to less than 4 mm [58, 59]. The time resolution of determining the mean ortho-positronium lifetime (τ*_oPs_*) scales inversely proportional to the square root of the number of identified oPs events [5, 9]). Hence, by extrapolating the result obtained in this study, we anticipate that the use of the high-sensitivity scanners mentioned above will enable τ*_oPs_* imaging with a temporal resolution better than 10 ps, which can potentially be further improved using iterative methods [11]. It is worth noting that a temporal resolution of 20 ps was already achieved in the first *ex vivo* positronium imaging [9], and that other methods for improving the precision of lifetime spectra analysis are actively being developed [11, 58–63].

The resolution of 10 ps is much smaller than the range of oPs lifetime values for various tissues, ranging from about 1.4 ns to 2.9 ns [10, 12, 25–27, 64]. It is also smaller than differences observed in the oPs mean lifetime between healthy and cancer tissues, ranging from 50 ps to 700 ps [9, 10, 13, 28, 65, 66]. Consequently, the advent of positronium imaging with a 10 ps resolution, which is achievable by total-body PET systems, may be useful for the in vivo assessment of tissue pathology. Such accuracy of oPs lifetime imaging could, in principle, enable the use of positronium as a biomarker of hypoxia to distinguish between hypoxic and normoxic states in tissue *in vivo*. The partial pressure of oxygen in cancerous tissues is approximately 10 to 50 mmHg lower than in healthy tissues, depending on the tissue type [24, 67]. The changes in oPs lifetime in the tissues of living organisms due to variations in oxygen pressure are not yet known. In water, these changes are very small, at the level of a few picoseconds [22–24]. However, in cells, they are expected to be on the order of 50 picoseconds, based on the analogy with the change in mean oPs lifetime in organic liquids [22, 23] such as isopropanol, which has a structure very similar to that of many typical cell metabolites. In isopropanol the mean oPs lifetime (via the conversion mechanism shown in Fig. 1) changes by about 50 ps when the oxygen pressure changes by 10 mmHg [22, 23].

Notably, results shown in Fig. 3N demonstrate that the determined mean positron annihilation lifetime exhibits statistically significant differences between glioma (1.28 *±* 0.06 ns), salivary glands (1.53 *±* 0.04 ns), and healthy brain tissues (1.89 *±* 0.06 ns). The mean positron lifetime denotes the average time between positron emission and annihilation, averaged over all possible annihilation mechanisms (illustrated in Fig. 1). Consequently, it can be interpreted as an effective parameter reflecting both intramolecular void size and oxygen concentration (primarily detected by ortho-positronium) and bulk molecular structure (primarily influencing direct annihilation). Although the mean positron lifetime is less sensitive to partial oxygen pressure than the oPs lifetime, its determination is simpler (merely averaging lifetimes without requiring deconvolution of the lifetime spectrum) and may prove more robust for clinical applications as a measure of bulk molecular alterations.

Lastly, it is noteworthy that positronium imaging provides complementary information about inter- and intramolecular structure and the degree of tissue oxidation compared to the information about metabolic rate (e.g., fluoro-deoxy-glucose uptake), anatomy (electron density), and morphology (hydrogen density) currently attainable with PET, computed tomography, and magnetic resonance tomography, respectively [9]. The diagnostic potential of positronium images remains to be fully explored by establishing an *in vivo* correlation between the lifetime of positronium and positrons in tissues and the degree of tissue alterations. Here, we take a first step in this direction by demonstrating positronium imaging of the human brain *in vivo*.

PET, with its unparalleled imaging sensitivity, has revolutionized the field of precision medicine [16, 30]. We envision that positronium imaging could transcend the current PET diagnostic paradigm, paving the way for *in vivo* tissue assessment at the sub-nanometer scale [1].

## IV. METHODS

### A. Description of the patient, bioethical committee

The imaging study involving the “Clinical application of the prototype J-PET apparatus” was performed at the Department of Nuclear Medicine of the Medical University of Warsaw in March 2022. The study was approved by the Ethical Committee at the Medical University of Warsaw (opinion from February 21, 2022 No. KB/16/2022). Informed consent was obtained from the patient to enter the study and to publish the results. The examined patient was a man in his 40s, diagnosed between 2011 and 2015 with a grade II primary brain glioma according to WHO CNS5 classification [68]. The tumor is located in the right frontoparietal lobe. In further histopathological examination performed in 2021, the brain tumor was classified as grade IV. The SUVmax value of 8.4 was determined in the study presented in this manuscript.

### B. Radiopharmaceutical preparation

^68^Ge/^68^Ga GalliaPharm generator, Eckert & Ziegler, Germany was used for all ^68^Ga labeling. Radio-pharmaceutical preparation of [^68^Ga]Ga-PSMA-11 is described in detail in reference [8] and [^68^Ga]Ga-DOTA-SP in reference [69]. The preparation of therapeutic doses of [^225^Ac]Ac-DOTA-SP is described in reference [29].

### C. Description of PET/CT

PET/CT examinations were performed using a Biograph 64 TruePoint PET/CT scanner (Siemens Medical Solutions, Knoxville, TN). The course of imaging is shown in Fig. 2A. Data were acquired in the 3D mode. CT data were used for the attenuation and scatter correction as provided by the vendor. The reconstruction was performed using the TrueX algorithm (Siemens Medical Solutions) with Point Spread Function (PSF) modeling, 3 iterations, and 21 subsets. Post-reconstruction 3D Gaussian filtering was performed with FWHM = 4 mm.

### D. Description of the modular J-PET tomograph

The modular J-PET scanner (Fig. 5), used in the presented research, is the first portable PET tomograph consisting of 24 independent detection modules, where each module is composed of 13 axially arranged plastic scintillator strips with dimensions of 24 × 6 × 500 mm^3^. A single module constitutes an independent detection unit comprised of scintillator strips coupled to the front-end electronics board that transfers digitized information about the registered signals to the Data Acquisition System (via optical links visible as orange cables in Fig. 1 and Fig. 2). The modular J-PET system, with visible scintillator strips and electronics boards, is shown in a photograph in Fig. 5A. The detection modules are lightweight (approximately 2 kg each), can be arranged in various configurations, and can be easily moved within the diagnostic room. For this research, they were arranged in a cylinder with an inner diameter of 74 cm and positioned behind the Biograph 64 TruePoin PET/CT scanner as can be seen in Fig. 2. The modular J-PET scanner and its performance, encompassing signal reconstruction, event reconstruction, and event selection procedures, are comprehensively detailed in reference [34]. For the sake of manuscript completeness, only a concise overview of the most important features is provided herein.

**Fig. 5:**
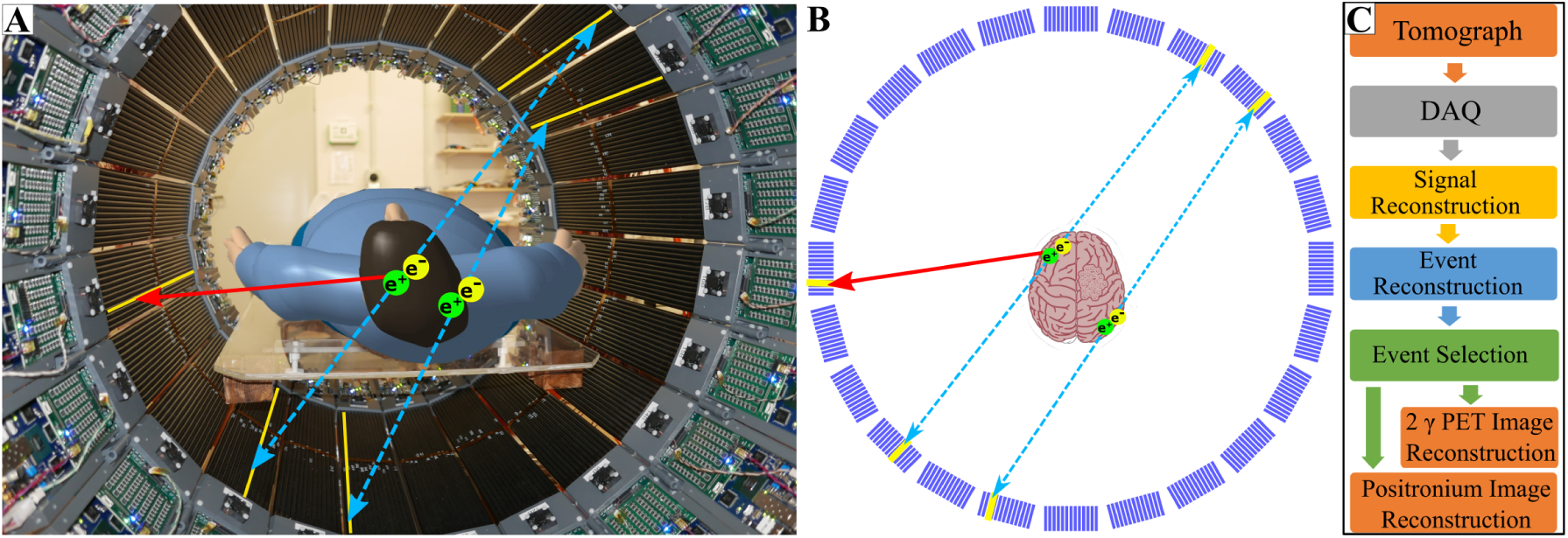
**(A)** Photograph of the modular J-PET system with a human placed inside the scanner. The superimposed blue-dashed arrows indicate photons from the electron-positron annihilation (e^+^e*^−^→* γγ), while the red-solid arrow indicates prompt gamma (γ*_p_*) from the decay of ^68^Ga radionuclide. Modular J-PET scanner enables registration and identification of both γγ and γγ+γ*_p_* events which are used in this study for the standard PET imaging (γγ), and positronium imaging (γγ+γ*_p_*). **(B)** Cross section of the modular J-PET showing the 24-module structure of the scanner, where each module is composed of 13 scintillator strips. Scintillators that register the photons are marked in yellow in Figs. A and B. **(C)** Block diagram of the data processing [36]. Light signals generated by gamma photons in the *Tomograph* are collected by a triggerless data acquisition system (*DAQ*) [35] and stored on discs in the form of the continuous sequence of timestamps. Time stamps are used for *Signal Reconstruction* including place and time of gamma photon interaction. Signals are then used for *Event Reconstruction*. Next, the true γγ and γγ+γ*_p_* events are selected by *Event Selection* algorithms. Finally, the selected events, in the list mode format, are used for the standard *2*γ *PET Image Reconstruction*, and *Positronium Image Reconstruction*.

Plastic scintillator strips (Saint Gobain BC-404) are wrapped in Vikuiti Enhanced Specular Reflector (ESR) foil to improve light propagation and DuPont Kapton 100B film to ensure light-tightness [32]. At the ends of each scintillator strip, on both 24 × 6 mm^2^ surfaces, 4 Hamamatsu-S13 Silicon Photomultipliers (SiPMs), with the area of 6 x 6 mm^2^, are connected optically by using Saint Gobain BC-600 Optical Cement. The gamma-ray emitted from the patient induces in the scintillator optical photons which are converted to electric signals by 8 SiPMs. The bias voltage at all SiPM is set to 56 V. SiPM signals are amplified (x13) and sampled at the two voltage levels of 30 mV and 70 mV (see Fig. 6C) using dedicated front-end electronic boards, described in detail in reference [33]. As a result of gamma photon interaction in a scintillator strip, two timestamps are recorded at the leading and two timestamps at the trailing edge on each signal from the eight SiMPs. Timestamps recorded by the front-end electronics are collected in the continuous mode (referred to as *triggerless* or *single interaction* mode) by the dedicated Data Acquisition System (DAQ) described briefly in the next section, while for details the reader is referred to reference [35]. The possibility of triggerless recording makes the modular J-PET the unique multi-photon scanner, enabling simultaneous detection of annihilation photons (2γ) and prompt gamma (γ*_p_*). Fig. 4A shows the sensitivity profile of the modular J-PET scanner for standard 2γ PET imaging (red), and for 2γ + γ*_p_* positronium imaging (green). At the center, the sensitivity for 2γ PET imaging amounts to 3.4 cps/kBq, while the sensitivity for 2γ + γ*_p_* imaging is equal to 0.06 cps/kBq.

**Fig. 6:**
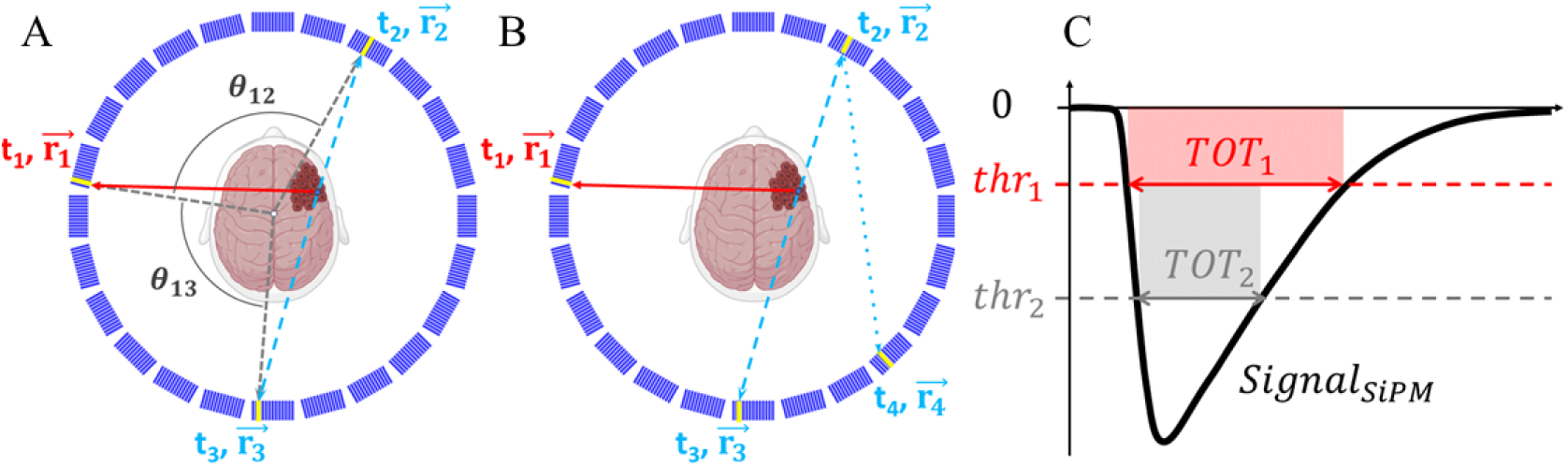
**(A)** Schematic cross-section of the modular J-PET scanner with the superimposed illustration of an exemplary event used for positronium imaging. A deexcitation photon and two annihilation photons are shown as solid-red line and dashed-blue lines, respectively. Scintillators that registered photons are marked in yellow. The reconstructed position and time of photons’ interaction (hit-position and hit-time) are indicated as 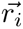 and t*_i_* (i = 1 .. 3), respectively. For the purpose of the event selection, the relative angles between photons (θ_12_, θ_13_) are calculated with respect to the center, as angles in the plane transverse to the scanner axis (XY plane), as illustrated in the figure. **(B)** An example of an event with more than three hits which are also used for positronium imaging. The position and time of interaction of the scattered photon is indicated as (t_4_, 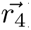). **(C)** Pictorial definition of the Time Over Threshold (TOT) for a single silicon photomultiplier (SiPM) signal. For each SiPM the time stamps at the leading and trailing edge of the signals are measured at two predefined voltage levels (thresholds) - thr_1_ and thr_2_. This enables us to determine the signal widths at two levels: TOT_1_ and TOT_2_, and to estimate the area of the signal as TOT*_SiPM_* = thr_1_ *∗* TOT_1_ + (thr_2_ *−* thr_1_) *∗* TOT_2_. The average of TOT*_SiPM_* from eight SiPMs attached to each scintillator is used as a measure of the deposited energy [70].

### E. Data Acquisition System

The Data Acquisition System is designed to register multi-photon events. Examples of such events useful for positronium imaging are shown in Fig. 6A and Fig. 6B. The complete J-PET DAQ system is based on Field-Programmable Gate Array (FPGA) electronics which allow for an efficient pipelined processing of multiple data streams in real-time [35]. Its architecture is hierarchical with the Controller board as the central point, Concentrator boards as hubs, and Endpoints that are digitizing boards located on the detector modules. The communication between the system components is implemented on optical links connections with Xilinx Aurora protocol running at 5 Gbps. A single link is bidirectional and shares three logical functionalities: transport of the measurement data, exchange of control and monitoring messages, and precise time synchronization of the Endpoints. The Controller board serves as an interface for the system operator to interact with all components in the system and generates the common synchronization signal that is propagated throughout the system components. The data readout is performed in continuous mode without any sort of hardware trigger. Digitized detector responses are stored in data buffers on the Endpoints and await the synchronization message to arrive. The buffer content is sent back to the Concentrators, which aggregate data from multiple Endpoints, encapsulate it into UDP packets, and transfer it out of the system to the storage using a 10 Gigabit Ethernet network. The applied data acquisition system is described in detail in reference [35].

### F. Signal Reconstruction

Raw data collected with the triggerless DAQ [35] are stored on discs as a sequence of timestamps determined relative to the synchronization time generated by the DAQ system every 50 µs. The offline processing of signals is conducted using the dedicated programs embedded in the J-PET Analysis Framework described in detail in reference [36].

The data processing involves assembling timestamps into representations of the SiPM signals and calculating the SiPM signal area (TOT*_SiPM_*) as explained in Fig. 6C. Next for each scintillator strip, the arrival times (t*_left_* and t*_right_*) of the light signal to the left and right scintillator end are calculated. The arrival times t*_left_* and t*_right_* are determined as an average of timestamps registered for the leading edge at the lower threshold for four SiPMs attached to the left and right side, respectively. Further on, if the difference between t*_left_* and t*_right_* times is smaller than 12 ns then they are considered to originate from the same gamma-ray interaction. The interaction time, referred to as the hit time, is estimated as t*_hit_* = (t*_left_* + t*_right_*)/2, while the interaction position along the scintillator strip with relative to the strip’s center is estimated as z*_hit_* = (t*_left_* - t*_right_*)*×* v*_s_*/2, where v*_s_* denotes the speed of light signal propagation in the scintillator strip. The method is described in detail in references [32, 71].

In the above-described estimations of interaction time and position, the corresponding calibration constants were applied individually for each scintillator strip, each SiPM, and each electronic channel. The calibration of all detector components was performed using point-like ^22^Na and ^44^Sc radioactive sources and applying methods described in detail in references [9, 33, 35, 71, 72].

The energy deposition in a scintillator strip is estimated based on the time-over-threshold value (TOT*_hit_*) using the relation established in reference [70], where TOT*_hit_* is calculated as an average of TOT*_SiPM_* values from SiPMs attached to the scintillator. Fig. 7A shows the TOT*_hit_* distribution reflecting contributions from continuous energy deposition via Compton scattering of (i) 511 keV annihilation photons, (ii) 1077 keV prompt gamma from ^68^Ga decay, (iii) photons resulting in scatterings in the detector, and from (iv) photons from decays of oPs and direct annihilations into three photons with energies lower than 511 keV. The continuous spectrum due to 511 keV photons ends (at the so-called *Compton edge*) at the TOT*_hit_* value of about 7 ns*·*V, while the spectrum due to the 1077 keV prompt gamma extends to higher TOT*_hit_* values marked in green. Prompt gammas are emitted in only about 3% of the ^68^Ga decays (see decay scheme in Fig. 4B). The TOT hit value is employed as a primary criterion to distinguish signals arising from annihilation photons (marked in red) and prompt gamma rays (marked in green).

**Fig. 7:**
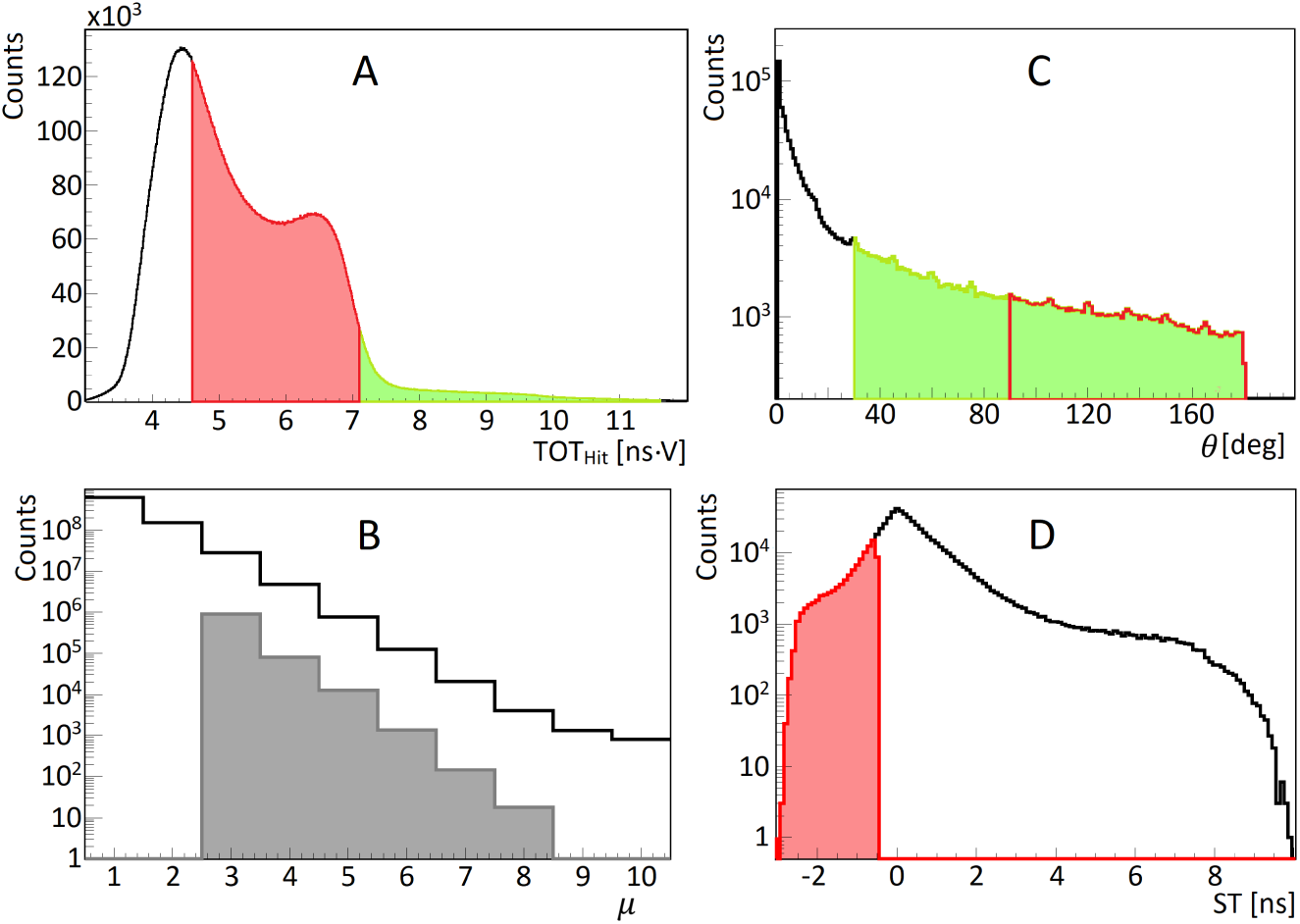
**(A)** Distribution of time-over-threshold (TOT*_Hit_*), used for photon identification. The ranges of TOT*_Hit_* values for selecting annihilation and deexcitation photons are marked in red and green, respectively. **(B)** Distribution of a number of hits in the event (hit multiplicity µ). The grey-shadowed histogram shows the hit multiplicity for events including one identified deexcitation photon, two identified annihilation photons, and other possible hits identified as scattered photons. **(C)** Distribution of relative angle (θ) between hit position vectors 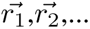 (defined in Fig. 6A). The figure includes all relative angles in the measured event (θ_12_, θ_23_, …). In the analysis, the relative angle between deexcitation and annihilation photons is restricted to the range marked in green (θ > 30*^o^*), and the red range (θ > 90*^o^*) shows the restriction used for the relative angle between annihilation photons. **(D)** Distribution of ST (*Scatter Test*) value used to suppress misidentification of scattered photons as annihilation photons (indicated in Fig. 8A). For hits assigned to annihilation photons (t_2_, 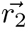) and (t_3_,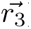) the ST is defined as a difference between the measured times (Δt = *|*t_3_ *−* t_2_*|*) and the time the photon would need to pass from 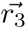 to 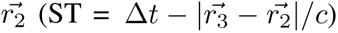. ST is equal to zero for the scattered photons, while for the annihilation photons, ST is negative. The region used to select events with annihilation photons (ST < 0.5 ns) is marked in red. Such selection reduces also a large fraction of events due to the accidental coincidences (as indicated in Fig. 8D) that are spread over the whole range of ST.

### G. Event reconstruction and selection

As an event, we define all processes in the detector system that are due to the single decay of radionuclide administered to the patient. Signals in scintillator strips (that reconstruction was described in section IV-F) are assumed to originate from the same event if their interaction times (t*_hit_*) are within 10 ns time window. Fig. 7B shows the distribution of the number of registered interactions per event (distribution of multiplicity of hits in the event). The solid-line histogram illustrates that, as anticipated, single-hit events are the most probable, and with each additional hit, the probability of event occurrence decreases by approximately a factor of five.

For positronium imaging, it is required to select events in which two hits are due to annihilation photons and one hit is due to the prompt gamma as indicated e.g. in Fig. 1A and Fig. 6A. Such selection is performed by requiring two hits with the TOT*_hit_* value in the red range defined in Fig. 1A, and one hit in the green range. The shadowed grey histogram in Fig. 7B shows the distribution of hit multiplicity after selecting such events. The multiplicity of hits larger than 3 corresponds to events when the annihilation or prompt photon interacts more than once in the detector. An example of such events is shown in Fig. 6B.

The next step of the data selection aims at the suppression of background events in which the scattered photon was misidentified as an annihilation photon or in which prompt gamma and annihilation photons originate from two different radionuclide decays. Examples of such events are shown in Fig. 8A-D. These background events are suppressed by restricting the relative angle θ*_ij_* between vectors pointing from the center to the i*^th^* and j*^th^* interaction points as defined in Fig. 6A. The experimental distribution of this angle (discussed also in detail in reference [70]) is presented in Fig. 7C. The relative angle between the annihilation photons and prompt gamma is distributed uniformly since they are emitted independently, while the background events with the prompt gamma scattering (Fig. 8A) are at most probable in the adjacent scintillators at small values of θ. Therefore, accepting only events with θ larger than 30*^◦^* (indicated by the green range in Fig. 7C) significantly (by a factor of four) reduces the background from prompt gamma scattering (Fig. 8A), while signal events are reduced only by about 17%. The remaining background from the scattering of the annihilation photons (Fig. 8B) and two types of accidental coincidences (Fig. 8C and Fig. 8D) was suppressed by further restrictions of the range of the angle θ, and by the application of the *scatter test (ST)* criterion defined in the caption of Fig. 7D. Restricting the angle θ between the annihilation photon candidates to values larger than 90*^◦^* (range marked in red in Fig.7C) reduces a large fraction of the background shown in Fig. 8B, since the scattering is at most probable in the adjacent scintillators. In addition, it reduces also about 50% of the accidental coincidence of the type shown in Fig. 8D. Notably, this criterion corresponds only to the reduction of the imaging field of view from 74 cm diameter to 52 cm diameter and so does not affect the useful signal events. The misidentified background events caused by the scattering of annihilation photons (Fig. 8B) and accidental coincidences (Fig. 8D) with an angle θ greater than 90*^◦^* are further suppressed using the scatter test as shown in Fig. 7D and explained in the figure caption.

**Fig. 8:**
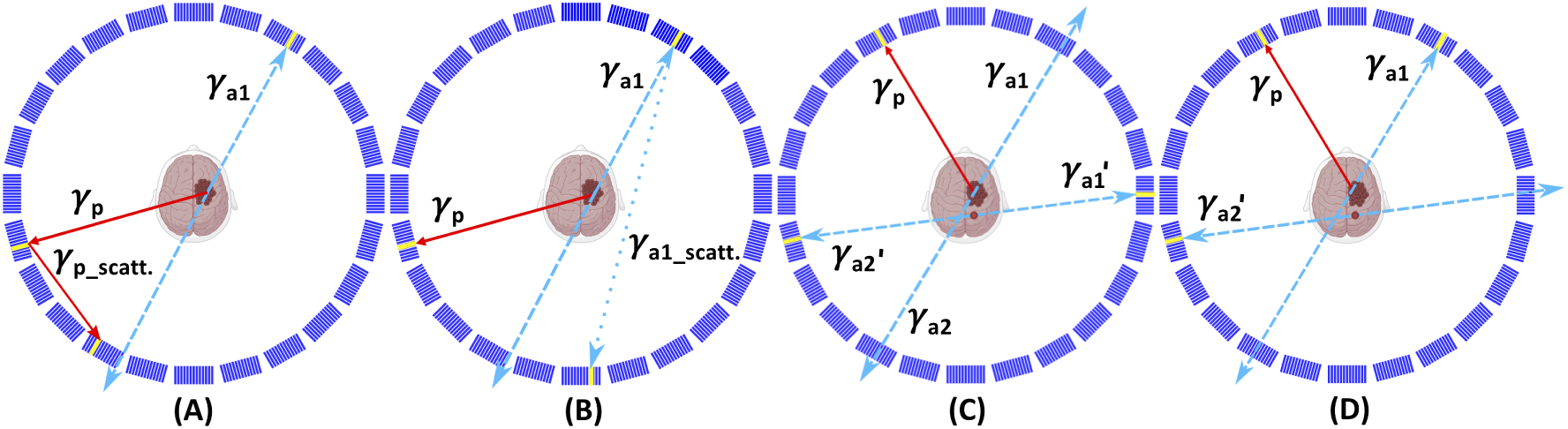
Examples of background events superimposed upon the cross-section of the modular J-PET scanner. The prompt gamma is represented by a solid red arrow. Primary and scattered annihilation photons are indicated by blue dashed and blue dotted arrows, respectively. Scintillators where photons interacted are highlighted in yellow. **(A)** An example of a background event where one annihilation photon remained undetected while the prompt gamma interacted twice within the detector, leading to the misidentification of the registered scattered photon as the annihilation photon. **(B)** An example of a background event where one annihilation photon remains undetected, while the other annihilation photon interacts twice within the detector, leading to the misidentification of the registered scattered photon as an annihilation photon. Such events can be suppressed using the scatter test (Figure 7D). **(C)** An example of background arising from the accidental coincidence of a prompt gamma from one event and annihilation photons from the other event. A portion of these events can be eliminated using the scatter test (Fig. 7D). **(D)** An example of background arising from the accidental coincidence of registering a prompt gamma and one annihilation photon from one event, and the other annihilation photon from the other event. A portion of these events can be eliminated by restricting the range of angle θ (Fig. 7C) and by using the scatter test (Figure 7D).

Events that met all the aforementioned selection criteria were employed for the positronium image reconstruction described in the subsequent section. The lifetime spectra depicted in Figure 3J-L demonstrate that the background under the signal (within the range of 0 to 5 ns) was reduced to a level of approximately 20%.

### H. Positronium image reconstruction

Selected events comprising hits from prompt gamma (t_1_, 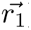) and from two annihilation photons ((t_2_, 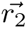) and (t_3_, 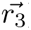)) are used to reconstruct positronium image by application of the method described in detail in reference [9]. In short, the positronium image reconstruction procedure is as follows. For each event separately, the annihilation place 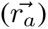 and the positron lifetime in the patient’s body (ΔT) are determined. The most probable position 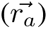 and time (t*_a_*) of positron-electron annihilation is calculated as:

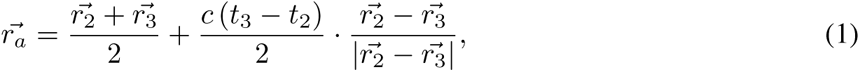

and

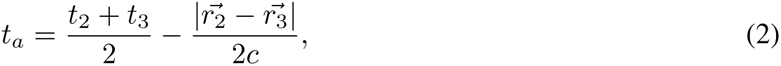

where c denotes the speed of light.

The positron lifetime (ΔT = t*_a_* - t*_p_*) is next calculated as a difference between annihilation time (t*_a_*) and the time of the positron emission. In the case of the ^68^Ga radionuclide (as illustrated in the decay scheme in Figure 4B), the prompt gamma is emitted, on average, only 1.6 ps after the emission of a positron. This extremely short delay can be safely disregarded. Therefore, the time of positron emission is approximated by the time of prompt gamma emission (t*_p_*). The emission time of the prompt gamma is determined by correcting the time of its registration by the time it took to travel from the emission point to the registration point 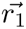:

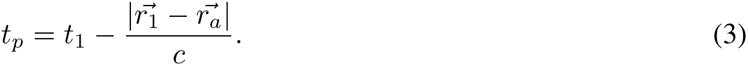

In the above calculations, the place of the prompt gamma emission was approximated by the place of positron annihilation. This is justified because in the case of positrons emitted by ^68^Ga radionuclide, the average distance in the tissue between the positron emission and annihilation is equal to about 2.3 mm [47], which is small with respect to the spatial resolution of determining 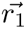. The 3D-tomography image of places of positron-electron annihilations accompanied by the emission of prompt gamma is determined by calculating the number of annihilation points in each voxel of the head of the patient. The transverse, coronal, and sagittal cross-sections of the reconstructed images are shown in Fig. 3D-F. The presented positronium image cross-sections were selected to clearly visualize annihilations in glioblastoma cancer. Due to the low number of events registered in this study, the patient head was divided only into three regions of interest including glioblastoma cancer (G), salivary glands (SG), and the rest constituting the healthy brain tissues (B). The chosen regions are large and include the scalp and other head tissues. However, due to the high affinity of PSMA and Substance-P to glioblastoma and salivary glands [8, 30], the signal registered may be well considered as originating predominantly from these tissues. The positron lifetime spectra from these regions of interest are shown in Fig. 3J-L, while the mean lifetime of oPs and the mean value of positron lifetime are presented in Fig. 3 M and N. The method of determining the mean lifetime of oPs in the regions of interest is described in the next section.

### I. Positronium mean lifetime determination

The determined positron lifetime spectra (shown as black histograms in Fig. 3J-L) were used to estimate the mean lifetime of ortho-positronium for each region of interest. In principle, the ΔT spectrum, determined for the tissue, comprises contributions from [2] (i) the direct electron-positron annihilations (*∼* 60%), (ii) annihilations intermediated by para-positronium (*∼* 10%), (iii) annihilations intermediated by ortho-positronium (*∼* 30%), and the background from accidental coincidences. All these contributions are smeared due to the experimental resolution. The ΔT resolution of the single measurement of ΔT was estimated to 520 ps (FWHM). The width of the bin in the ΔT spectra is equal to 0.4 ns, which is the value close to the FWHM of the ΔT resolution.

In the first step of the positron lifetime spectrum analysis, the constant background from accidental coincidences was estimated by taking the mean value in the ΔT range: −10 ns < ΔT < −5 ns. In this range of negative ΔT values, only events caused by random coincidences are expected. The obtained values for the background contributions (expressed in counts per bin) are equal to 8.5 *±* 0.9 for glioblastoma, 13.6 *±* 1.1 for salivary gland, and 11.9 *±* 1.1 for the healthy brain. The background contributions are indicated as violet-dotted lines in Fig. 3J-L. Next, in order to estimate the mean oPs lifetime, the sum of exponential contributions convoluted with the experimental resolution was fitted to the experimental spectrum using a dedicated PALS Avalanche software, described in detail in references [37–39]. Due to the low statistics of events and the time resolution being several times larger than the average pPs lifetime, only the mean oPs lifetime was considered as a free parameter to ensure the stability of the results. The contributions from pPs and direct annihilations were considered as a single component with a fixed mean lifetime of 0.329 ns and 70% intensity (cyan line in Fig. 3J-L). These values were determined based on previous *ex vivo* studies of positronium lifetime in the tissues [9]. The oPs mean lifetimes obtained from the fit are presented in Fig. 3M. In addition, the mean value of ΔT in the signal region between 0 and 5 ns was calculated. The mean was calculated after subtracting the background. The resulting values of the mean positron lifetime in the studied region of interest are shown in Fig. 3N.

### J. Monte-Carlo simulations of sensitivity profiles of the PET scanners

The sensitivity profiles and gains presented in Fig. 4 have been determined using the GATE software version 9.0[73] extended by the J-PET group (https://github.com/JPETTomography/Gate). GATE (Geant4 Application for Tomographic Emission) is an advanced open-source software (www.opengatecollaboration.org/) based on the Geant4 toolkit [74] and dedicated to Monte Carlo simulations in fields of medical imaging and radiotherapy. In PET imaging, GATE allows for the generation and tracking of radioactive source decays and their products, simulation of the sensitivity to the detection scanners, and data processing. In this study, the emlivermore_polar was the utilized physics list. The materials and geometries of simulated scanners are described below. The Modular J-PET scanner consists of 24 modules arranged into single layer ring with a 740 mm inner diameter, where each module is composed of 13 axially arranged plastic scintillator strips with dimensions of 24*×*6*×*500 mm^3^. The Total-Body J-PET consists of 7 rings with an 830 mm inner diameter, 330 mm axial length and a 20 mm gap in between the rings. The total axial field of view was equal to 243 cm. Each ring includes 24 modules, where each module comprises two layers in radial direction formed from 16 plastic scintillator strips [40] with dimensions of 30*×*6*×*330 mm^3^.

As a material representing the plastic scintillator, the EJ-230 was chosen. For positronium imaging, the simulation included 511 keV annihilation photons and prompt gamma rays with an energy of 1160 keV. An energy deposition between 200 keV and 350 keV was required for annihilation photons, while an energy deposition greater than 350 keV was required for prompt gamma rays. The coincidence window was set to 4 ns for the modular J-PET scanner and to 4.5 ns for the total-body J-PET scanner.

The crystal-based scanners were simulated using LYSO crystals with a thickness of 1.81 cm and a geometrical arrangement similar to that of the uEXPLORER scanner [18]. The calculations were performed for different numbers of rings, including 2, 3, 4, 6, 8, and 10. For the crystal-based scanners, the coincidence time window was set to 4.5 ns. The energy window for annihilation photons was set to 430-645 keV, while for prompt gamma photons, an energy deposition greater than 645 keV was required.

## Data Availability

All data produced in the present study are available upon reasonable request to the authors

## Acknowledgements

The authors acknowledge the strong support of E. Beyene, A. Coussat, A. Heczko, H. Giemza, P. Kapusta, P. Konieczka, W. Migda}, S. Moyo and P. Wasiuk. The authors acknowledge also the support provided by the Foundation for Polish Science through the TEAM POIR.04.04.00-00-4204/17 program; the National Science Centre of Poland through grants MAESTRO no. 2021/42/A/ST2/00423 and OPUS no. 2021/43/B/ST2/02150; the Ministry of Education and Science through grant no. SPUB/SP/490528/2021; the SciMat and qLife Priority Research Areas budget under the program *Excellence Initiative - Research University* at the Jagiellonian University, and Jagiellonian University project no. CRP/0641.221.2020.

## Author contributions

The portable modular J-PET scanner and the positronium imaging method were conceived by P. M. P. M. and E. S. planned and supervised the research and interpreted the obtained results. The manuscript was prepared by P. M. in collaboration with E.S. and the input from J. B., K. D., A. G., G. K., D. K., Kubat, K. Kacprzak, S. N., S. P., S. S., L. Królicki, J. K., and was then edited and approved by all authors. Figures were conceptualized by P. M. and prepared by J. B., K. D., D. K., S. P., Shivani, and S. S. The performed medical experiment was planned by P. M, L. Królicki, and E. S. The ethical committee consent was secured by L. Królicki and J. K. The preparation of pharmaceuticals, planning of patient examination and imaging protocols, patient preparation, imaging, and therapy management were done by L. Królicki, J. K., K. F., M. K., and J. M. The imaging was conducted using the dedicated modular and portable J-PET apparatus developed by the J-PET collaboration. P. M., J. B., N. C., M. Das, K. D., A. G., G. K., W. K., D. K., W. M., S. N., S. P., L. Królicki, and E. S. installed the J-PET scanner in the hospital, put it into operation, and carried out imaging of the patient using J-PET. Preparation of ^44^Sc for the calibration of the detector was provided by J. Ch. The J-PET data analysis and calibrations were performed by K. Kacprzak, A. G., and K. D. under the supervision of P. M. Biograph PET/CT images were processed by J. B. Positronium lifetime images and their merging with CT and PET images were conceptualized by P. M. and performed by K. D. and J. B. under the supervision of P. M. Signal selection criteria were developed by P. M. Authors: P. M., J. B., N. C., C. C., E. C., M. Dadgar, M. Das, K. D., K. E., A. G., K. Kacprzak., M. Kajetanowicz, T. Kaplanoglu, Ł. Kapłon, G. K., T. Kozik, W. K., K. Kubat, D. K., W. Migdał, G. M., W. Mrykal, S. N., S. P., E. P.d.R., L. R., S. S., S., R.Y. S., M. Silarski, M. Skurzok, E. Ł. S., P. T., F. T.A., K. T.A., W. W., and E. S. participated in the construction, commissioning, and calibration of the modular J-PET scanner. S. N. and G. K. optimized the working parameters of the scanner. K. D., A. G., K. Kacprzak, and W. K. developed the J-PET analysis and simulation framework. K. Kacprzak and A. G. developed low-level signal reconstruction methods. Ł. Kapłon developed the methods and performed plastic scintillator characterization for the scanner construction. G. K. developed, programmed, synchronized, and operated the triggerless DAQ system. M. Kajetanowicz developed the front-end electronic boards for the scanner. W. Migdał assembled the J-PET detection modules and scanner mechanical constructions. K. Kacprzak and A. G. performed timing calibration of the detector. F. T. and S. N. determined NEMA performance characteristics of the modular J-PET scanner. K. Klimaszewski, W. W., L. R. and R.Y. S. managed the computing resources for high-level analysis and simulations. E. C. developed and operated short- and long-term data archiving systems and the computer center of J-PET. K. D. developed the PALS Avalanche program for the decomposition of positron annihilation lifetime spectra. S. S. established the relationship between energy deposition and TOT and the dependence of detection efficiency on energy deposition. S. P. performed Monte-Carlo simulations of sensitivity profiles. S. B. contributed with the theoretical support for positronium physics. P. M. and E. S managed the whole project and secured the main financing.

## Data and materials availability

All data needed to evaluate the conclusions in the paper are presented in the paper. The datasets collected in the experiment and analyzed during the current study are available from the corresponding author upon reasonable request.

